# SUMMIT: An integrative approach for better transcriptomic data imputation improves causal gene identification

**DOI:** 10.1101/2021.12.09.21267570

**Authors:** Zichen Zhang, Ye Eun Bae, Jonathan R. Bradley, Lang Wu, Chong Wu

## Abstract

Genes with moderate to low expression heritability may explain a large proportion of complex trait heritability, but these genes are insufficiently captured in transcriptome-wide association studies (TWAS) partly due to the relatively small available reference datasets for developing expression genetic prediction models to capture the moderate to low genetically regulated components of gene expression. Here, we introduce a new method, Summary-level Unified Method for Modeling Integrated Transcriptome (SUMMIT), to improve the expression prediction model accuracy and the power of TWAS by using a large expression quantitative trait loci (eQTL) summary-level dataset. We applied SUMMIT to the eQTL summary-level data provided by the eQTLGen consortium, which involve 31,684 blood samples from 37 cohorts. Through simulation studies and analyses of GWAS summary statistics for 24 complex traits, we show that SUMMIT substantially improves the accuracy of expression prediction in blood, successfully builds expression prediction models for genes with low expression heritability, and achieves higher statistical power than several benchmark methods. In the end, we conducted a case study of COVID-19 severity with SUMMIT and identified 11 likely causal genes associated with COVID-19 severity.

## 1 Introduction

Genome-wide association studies (GWASs) have shown that most disease-associated variants reside in non-coding regions [8, 25, 41], raising challenges in biological interpretation and target gene identification and validation [4]. These findings also lead to the hypothesis that genetic variants affect complex traits mainly through regulating gene expression levels, which motivates large-scale expression quantitative trait loci (eQTL) analyses [6, 33] and transcriptome-wide association studies (TWASs) [10, 11, 14, 27, 39, 44]. TWASs integrate expression reference panels (eQTL studies with matched individual-level expression and genetic data) with complex traits GWAS results to discover gene-trait associations. First, the expression reference panel is used to learn per-gene expression prediction model by regressing assayed gene expression levels on *cis*-eQTL genotypes (i.e., SNPs within 1 megabase of the gene transcription start site and transcription end site). Second, statistical associations are estimated between predicted gene expression levels for GWAS samples and the trait of interest. TWASs have garnered substantial interest within the human genetics community and have deepened our understanding of genetic regulation in many complex traits [12, 30].

Despite many successes, the size of expression reference panels primarily determines the number of analyzable genes, and hence the power of TWAS. For example, building expression prediction models with the Genotype-Tissue Expression (GTEx) project v7p data yielded more than twice more predictive models (i.e., analyzable genes) than that with GTEx v6p data (see TWAS/FUSION website, http://gusevlab.org/projects/fusion/gtex.html). For whole blood tissue, the number of analyzable genes increased from 2, 057 to 6, 006 when the size of the expression reference panel increased from 338 samples to 369 samples. Others have also observed that the number of analyzable genes can be significantly increased when using a slightly larger expression reference panel [44]. More importantly, perhaps due to the small sample size of available expression reference panels, the current standard practice of TWAS is to only analyze genes with model performance *R*^2^ ≥ 0.01 [10, 11, 14]. This practice may ignore genes with low expression heritability, but larger causal effect sizes on the trait of interest, since genes with low expression heritability have substantially larger causal effect sizes on complex traits [41]. It is of great interest to construct more powerful gene expression prediction models, especially for those genes with low expression heritability.

One potential approach to improve the power of TWAS is to directly combine individual-level reference panel data from several consortia or studies, increasing the sample size of expression reference panel. While straightforward, privacy concerns and subject consents can preclude access to individual-level reference panel data, making this approach often practically infeasible. On the other hand, one may use summary-level expression panels (often publicly available) with much larger sample sizes to build expression prediction models. However, to date, there is limited exploration regarding how one can build expression prediction models using a summary-level expression panel.

In this work, we introduce Summary-level Unified Method for Modeling Integrated Transcriptome (SUMMIT), a novel method that integrates summary-level expression reference panel data, derived from much larger sample sizes, with trait GWAS results to identify associated genes for the trait of interest. Specifically, we build gene expression prediction models in blood based on the eQTL summary-level data provided by the eQTLGen consortium [33]. To date, eQTLGen consortium conducted the largest meta-analysis involving 31, 684 blood samples from 37 cohorts [33], and the corresponding eQTL summary-level data have been released to be publically available. Through extensive simulation studies and analyses of GWAS summary statistics from 25 complex traits, we show that SUMMIT substantially improves the accuracy of expression prediction in blood, successfully builds expression prediction models for genes with low expression heritability, and identifies way more risk genes than benchmark methods. In the end, we conduct a case study on COVID-19 severity and identify 11 likely causal genes. The real data results are deposited into a user-friendly and freely available web portal (https://chongwulab.shinyapps.io/SUMMIT-app/), enabling practitioners to easily search and download significant gene-trait associations.

## 2 Results

### 2.1 SUMMIT overview

We develop SUMMIT, which extends the conventional TWAS methods [10, 11, 14, 27, 39], by leveraging eQTL summary-level data to predict expression levels. SUMMIT consists of three main steps. First, for each gene in the genome, we train expression prediction models using a penalized regression framework with eQTL summary-level data (e.g., eQTLGen [*N* = 31, 684; 33]). Next, we test associations between predicted gene expression levels and the trait of interest for each fitted expression prediction model with satisfactory performance (with *R*^2^ ≥ 0.005). Finally, we apply the Cauchy combination test to aggregate results from the fitted prediction models, which effectively quantifies the overall gene-trait associations.

### 2.2 Simulation results

In the simulation studies, we first evaluated the accuracy of expression imputation models generated by SUMMIT and benchmark methods and the corresponding statistical power. Next, we studied the impact of sample size on prediction performance and verified that SUMMIT recovered the information of individual-level expression reference panel from summary-level data.

First, we observed that SUMMIT performed better than two widely-used competing methods, TWAS-fusion and PrediXcan, yielding higher average imputation *R*^2^ with respect to different gene expression heritability 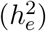 and the proportion of causal SNPs (*p*_causal_) (Figure 2a). For example, when 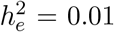 and *p*_causal_ = 0.2, the average imputation *R*^2^ of 1, 000 replications was estimated at 0.61% by SUMMIT, showing 1233% improvement compared with PrediXcan and 250% improvement compared with TWAS-fusion. Importantly, such improvements in expression prediction models result in consistently higher power of subsequent association studies under different sparsity levels (Figure 2b). For example, when 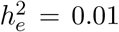 and *p*_causal_ = 0.2, the power of SUMMIT was 0.918 while that of PrediXcan and TWAS-fusion were 0.039 and 0.209, respectively.

**Figure 1:**
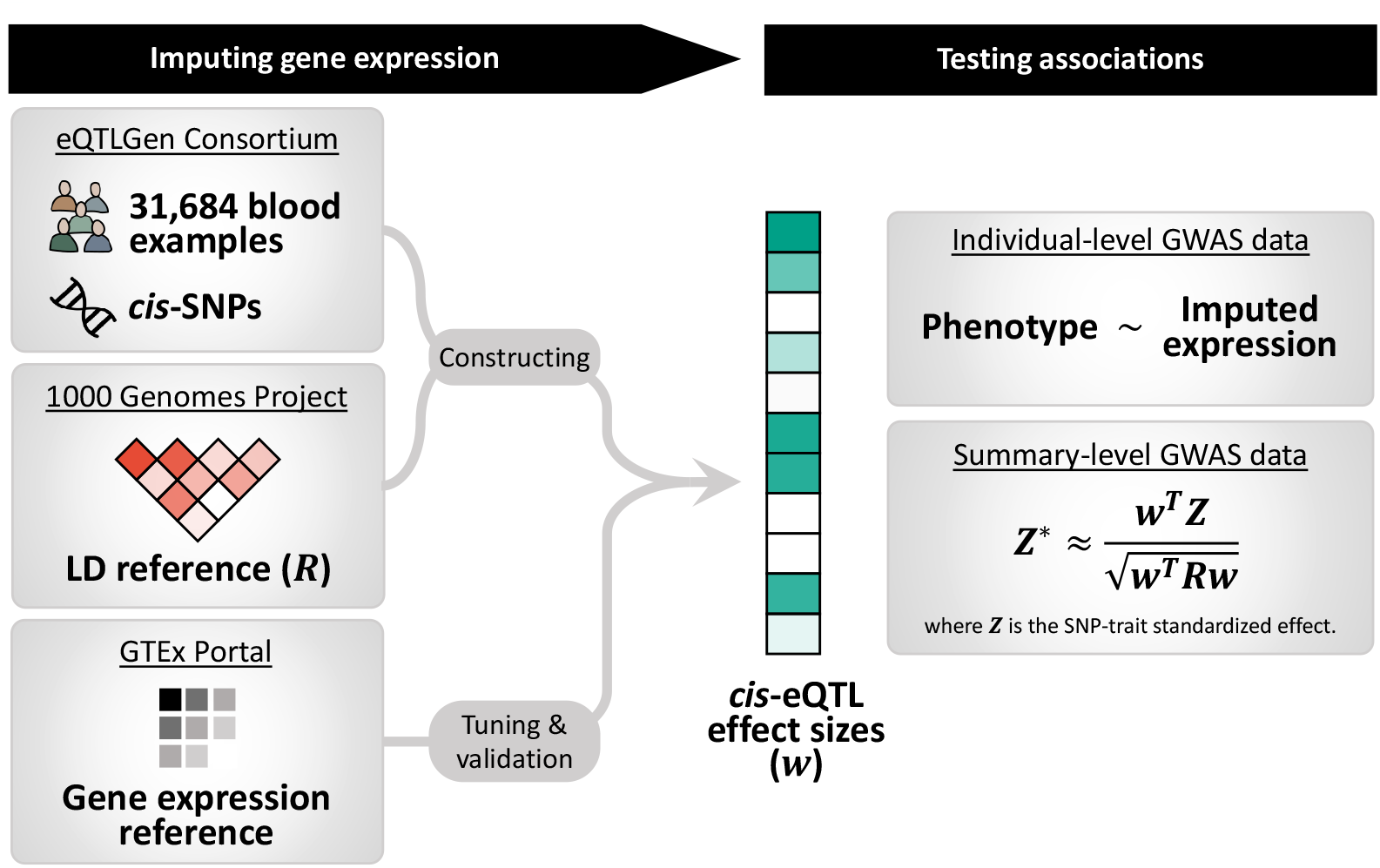
SUMMIT workflow. SUMMIT consists of three main steps: 1) building prediction models to impute gene expression levels; 2) testing association between predicted gene expression levels and the trait; and 3) aggregating results from all the fitted prediction models.

**Figure 2:**
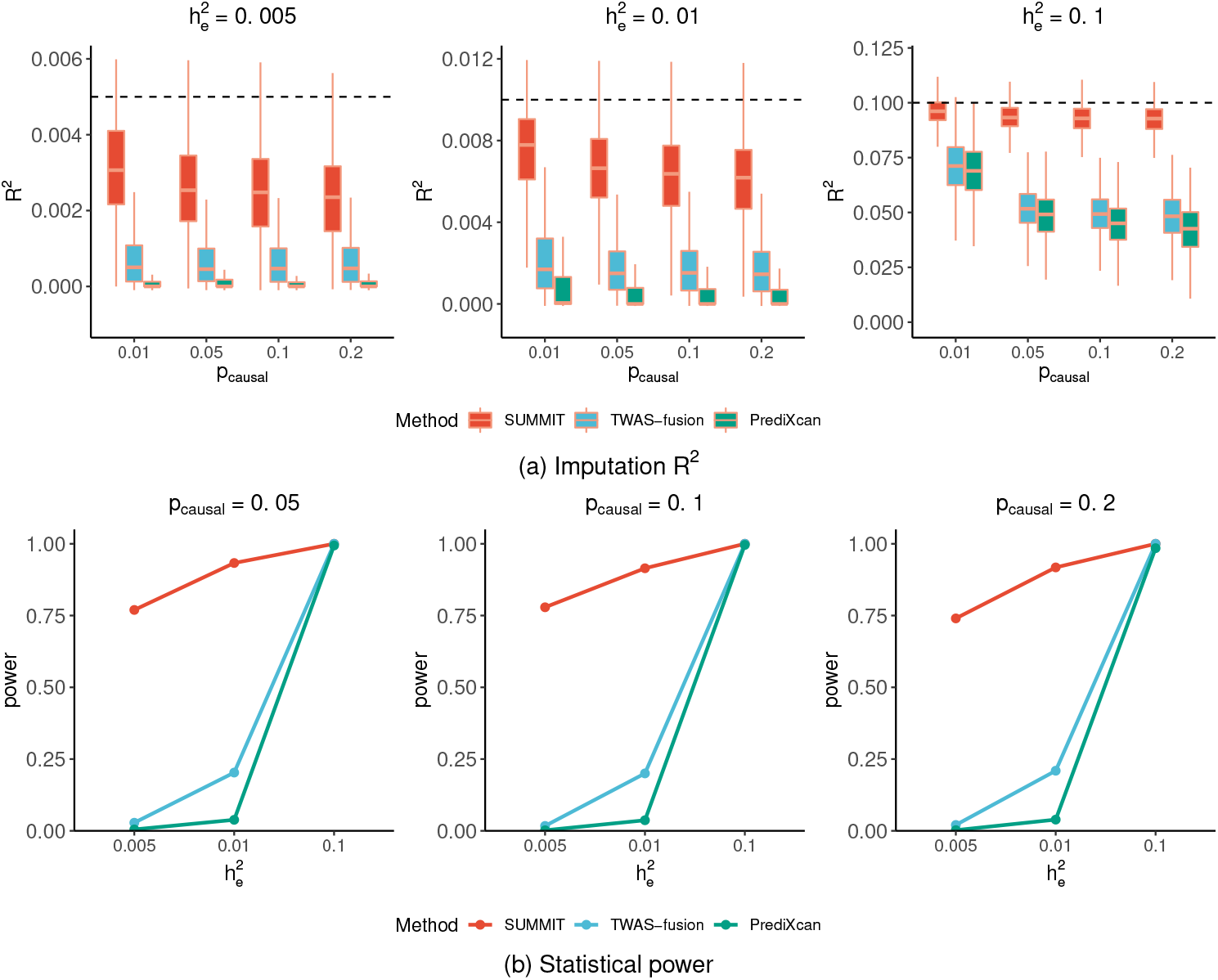
Performance comparison in simulations based on gene *CHURC1*. Plots of imputation *R*^2^ (a) and subsequent power (b) in test samples by SUMMIT, PrediXcan, and TWAS, with varying true expression heritability 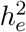 and proportion of true causal SNPs *p*_causal_. For (b), we set 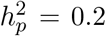 and empirical power was estimated by the proportions of *P*-value less than the significance threshold 2.5 × 10^−6^. The empirical power comparisons for 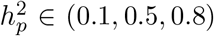 are in Supplementary Figure 1.

The current standard practice of TWAS is to only analyze genes with imputation *R*^2^ ≥ 0.01 and does not take into account genes with lower prediction performance (i.e., genes with imputation *R*^2^ between 0.005 and 0.01). However, such genes may have larger causal effect sizes on the trait of interest [41]. To evaluate the performance of different methods under highly small heritable situations, we simulated data with 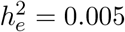. Figure 2a shows that SUMMIT achieved satisfactory performance under these scenarios. For example, when 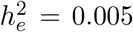 and *p*_causal_ = 0.2, SUMMIT estimated the average imputation *R*^2^ at 0.24%, which was much higher than those yielded by TWAS-fusion (0.067%; 255% improvement) and PrediXcan (0.014%; 1651% improvement). This is because SUMMIT leverages the summary-level eQTL data with a larger sample size.

Next, we studied the impact of the size of expression reference panel (Supplementary Figure 2). As expected, the imputation *R*^2^ increased as the sample size increased. For example, for the setting with 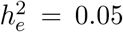 and *p*_*causal*_ = 0.2, when the sample size increased from 300 to 31, 684, the average imputation *R*^2^ increased from 0 to 0.0401, highlighting the gains of using a larger expression reference panel. Importantly, the imputation models became more stable (i.e., the variance decreased) as the sample size increased. Also, we confirmed that the imputation results from SUMMIT (average imputation *R*^2^: 0.0401) were highly similar to that of individual-level data available (average imputation *R*^2^: 0.0392), validating that SUMMIT can recover the individual-level information from summary-level data.

To consider the potential impact of genetic architecture, we considered two additional randomly selected genes, and the results were similar (Supplementary Figures 3–6). Furthermore, we ran simulations 5,000,000 times (5,000 runs for each of 1,000 computed weights) under the null hypothesis to evaluate the Type 1 error rates, confirming that all methods maintained well-controlled Type 1 error rates (Supplementary Figure 7).

In summary, these results demonstrated the potential usefulness of the SUMMIT for building expression prediction models and conducting subsequent association studies, especially for genes with low heritable expression.

### 2.3 SUMMIT improves the expression imputation accuracy

We compared the accuracy of expression imputation models of SUMMIT and four benchmark methods, including MR-JTI, TWAS-fusion, PrediXcan, and UTMOST for whole blood tissue. We trained the SUMMIT models with eQTLGen summary data, and the four benchmark methods were trained with GTEx data. For a fair comparison, we compared the number of genes with an estimated *R*^2^ ≥ 0.01 and only focused on the genes that appear in the eQTLGen summary data. The *R*^2^ for the benchmark methods were based on cross validation and provided by the corresponding authors, and the *R*^2^ for the SUMMIT was calculated based on the additional subjects in GTEx version 8 data, who have not been meta-analyzed in the eQTLGen and thus can be viewed as an independent external dataset. Compared with the benchmark methods, MR-JTI (9, 576 genes), TWAS-fusion (5, 411 genes), PrediXcan (7, 512 genes), and UTMOST (7, 236 genes), SUMMIT developed satisfactory prediction models for more genes (9, 749 genes with *R*^2^ ≥ 0.01). Importantly, SUMMIT could build prediction models for the majority (8, 364 out of 11, 450; 73%) of genes that can be analyzed by either of benchmark methods, (Figure 3(a)). In addition, SUMMIT was able to establish prediction models of additional 1, 836 genes that were ignored by benchmark methods, showing consistent improvement by using a large training dataset. Furthermore, compared with MR-JTI, SUMMIT achieved significantly higher prediction accuracy in different quantiles (*p* < 2.2 × 10^−16^ by the Kolmogorov-Smirnov test). This was also true for other methods (PrediXcan: *p* < 2.2 × 10^−16^; TWAS-fusion: *p* < 2.2 × 10^−16^; and UTMOST: *p* < 2.2 × 10^−16^).

**Figure 3:**
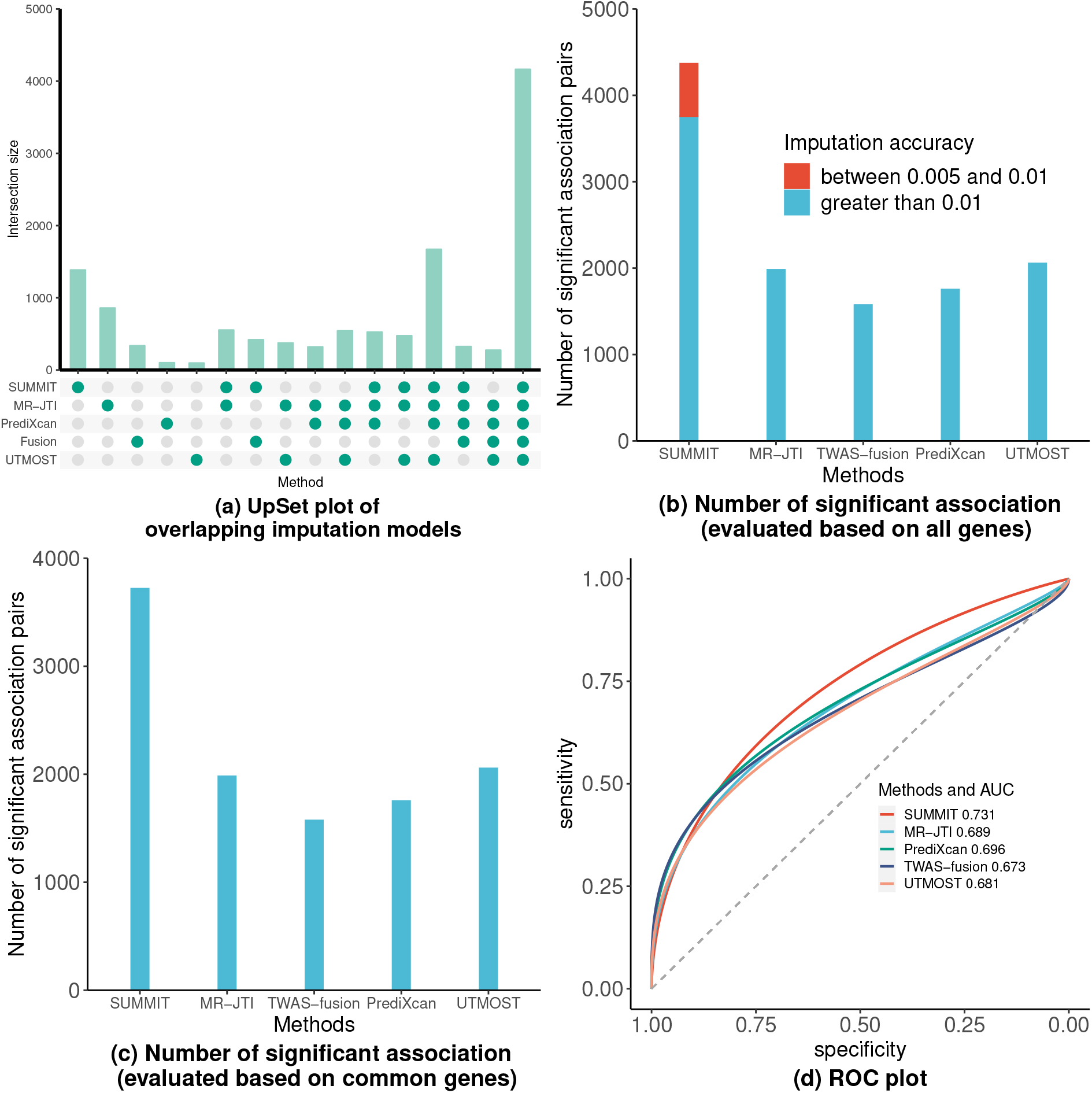
SUMMIT improves the performance of TWAS in real data. (a) is the UpSet plot of overlapping imputation models with *R*^2^ ≥ 0.01 among different methods. (b) shows the number of significant genes identified by different methods when using all available genes across 24 GWAS, where (c) shows the number of significant genes when evaluated on a common gene set of all methods. (d) is the ROC plot for identifying “silver standard” genes.

### 2.4 SUMMIT identifies more associations than competing methods

To evaluate the performance of identifying significant associations, we applied SUMMIT to the summary statistics of 24 GWAS (*N*_total_ ≈ 5, 600, 000 without adjusting for sample overlap across studies, Supplementary Table 1) and compared the results with those of the benchmark methods (for all genes with *R*^2^ ≥ 0.01). The full association results for SUMMIT were summarized in Supplementary Table 1. While the SUMMIT analyzed all genes with *R*^2^ ≥ 0.005 and applied the Bonferroni correction accordingly, we focused on the genes with *R*^2^ ≥ 0.01 for a fair comparison (Figure 3(b)). Compared with the benchmark methods, SUMMIT identified substantially more associations for each trait analyzed, showing 90% improvement compared with MR-JTI (*p* = 1.44 × 10^−5^ by the paired Wilcoxon rank test), 139% improvement compared with TWAS-fusion (*p* = 9.68×10^−6^), 115% improvement compared with PrediXcan (*p* = 5.96×10^−8^), and 83% improvement compared with UTMOST (*p* = 1.65 × 10^−5^).

Because different methods test different sets of genes, we also compared methods over a common set of 4, 291 genes that could be analyzed by all the methods (Figure 3(c)). Again, SUMMIT maintained an edge over competing methods, showing 35% improvement (*p* = 0.00053 by the paired Wilcoxon rank test) compared with the second best performing method, UTMOST.

Importantly, SUMMIT was applicable to analyze genes with low heritability for the expressions (0.005 ≤ *R*^2^ < 0.01), which have been largely ignored by benchmark methods. Out of the 11, 585 genes with *R*^2^ ≥ 0.005, 1, 836 had testing *R*^2^ between 0.005 and 0.01. For these 1, 836 genes, we identified 607 gene-trait associations (Figure 3(b)). In comparison, for the remaining 9, 749 genes, we identified 3, 759 gene-trait associations, indicating that genes with relatively smaller *R*^2^ may be equally important as those with larger *R*^2^. This finding is aligned with the fact that genes with low expression heritability have substantially larger causal effect sizes on complex traits [41].

### 2.5 SUMMIT achieves higher predictive power for identifying “silver standard” genes

We compared different methods in terms of identifying the likely causal genes that mediate the association between GWAS loci and the traits of interest. Following [2], we used a set of 1,258 likely causal gene-trait pairs curated by using the OMIM (Online Mendelian Inheritance in Man) database [13] and a set of 29 gene-trait pairs based on rare variant results from exome-wide association studies [18, 22, 24], which provide orthogonal information that is independent of GWAS results. Both gene-trait pairs sets can be found in Supplementary Table 2.

Figure 3(d) shows that SUMMIT yielded good sensitivity and specificity for identifying the silver standard genes and achieved the highest AUC (0.731) among all the methods compared. All methods achieved relatively good sensitivity and specificity, showcasing the potential predictive ability of TWAS-type methods to prioritize putative causal genes. For example, with Bonferroni correction cutoffs, SUMMIT identified 80 (54%) genes in the silver standard gene list, whereas the second best performing method, PrediXcan, identified 57 (39%). In summary, perhaps due to the improvement in expression prediction models, SUMMIT achieved higher predictive power in terms of prioritizing likely causal genes.

### 2.6 SUMMIT identifies novel risk genes for COVID-19 severity

We leveraged GWAS summary data from The COVID-19 HGI [16] to identify risk genes for COVID-19 severity. Using SUMMIT, we identified significant associations of 17 genes with COVID-19 severity (B2 outcome) comparing COVID-19 hospitalized patients and controls at the Bonferroni correction cutoff of 4.33 × 10^−6^ (Figure 4). In comparison, the competing methods PrediXcan, TWAS-fusion, UTMOST, and MR-JTI identified 1, 6, 2, and 1 significant genes, respectively (Supplementary Table 3). For these 17 genes identified by SUMMIT, 11 was prioritized by a fine-mapping method FOGS (Table 1). We further validated these 11 genes by analyzing COVID-19 comparing very severe respiratory confirmed COVID-19 versus population controls (A2). Of them, 10 were validated at *P* < 0.05.

**Figure 4:**
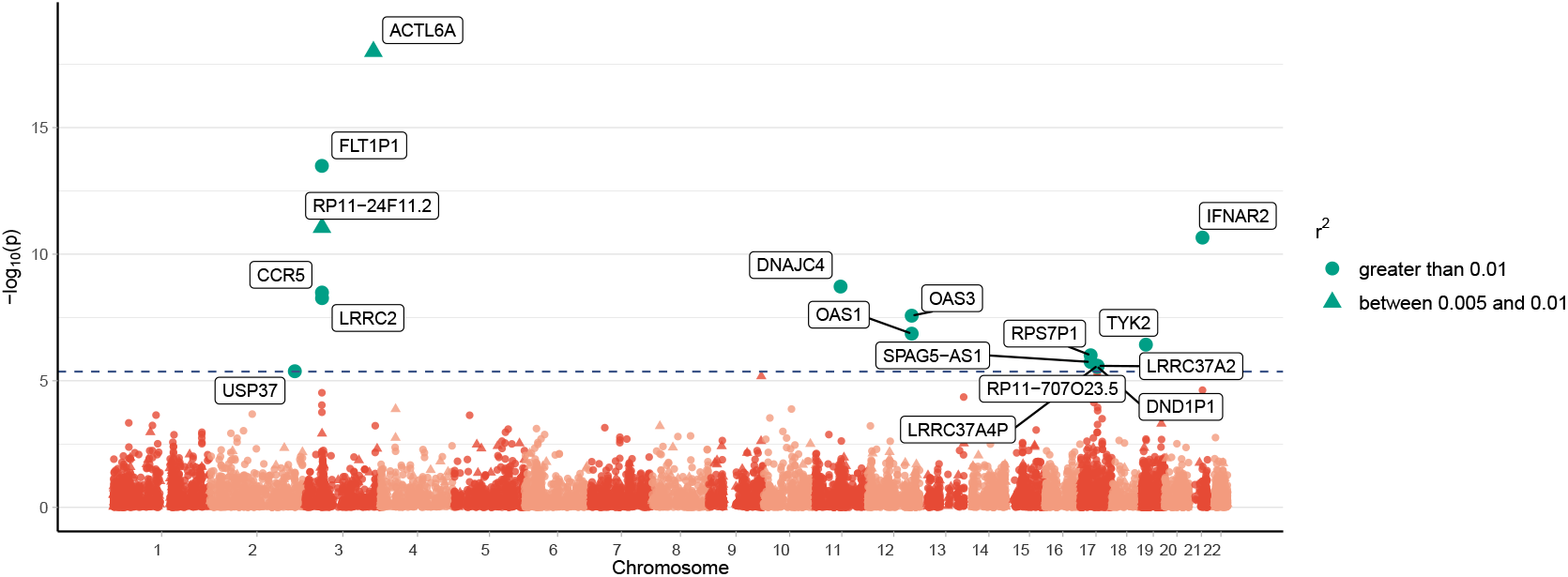
Manhattan plot for COVID-19 severity (B2 outcome) comparing COVID-19 hospitalized patients and controls. The horizontal line marks the genome-wide significance threshold (0.05*/*11539 ≈ 4.33 × 10^−6^).

**Table 1:** Predicted gene expression in blood–COVID-19 associations for the likely causal genes based on the COVID-19 Host Genetics Initiative data. “+” and “−” stand for positive and negative directions, respectively.

| Chromosome | Gene | $R^2$ | COVID-B2 | | COVID-A2 | |
| --- | --- | --- | --- | --- | --- | --- |
| | | | Direction | $p$ | Direction | $p$ |
| 3 | *ACTL6A* | 0.008 | — | $9.9 \times 10^{-19}$ | — | $2.5 \times 10^{-1}$ |
| 3 | *LRRC2* | 0.044 | — | $5.4 \times 10^{-9}$ | — | $3.9 \times 10^{-5}$ |
| 3 | *RP11-24F11.2* | 0.006 | — | $8.8 \times 10^{-12}$ | — | $9.0 \times 10^{-6}$ |
| 3 | *FLT1P1* | 0.116 | — | $3.3 \times 10^{-14}$ | — | $1.0 \times 10^{-7}$ |
| 3 | *CCR5* | 0.049 | — | $9.9 \times 10^{-19}$ | — | $1.8 \times 10^{-6}$ |
| 12 | *OAS1* | 0.056 | — | $1.4 \times 10^{-7}$ | — | $1.2 \times 10^{-8}$ |
| 12 | *OAS3* | 0.041 | + | $2.7 \times 10^{-8}$ | + | $3.4 \times 10^{-11}$ |
| 17 | *LRRC37A4P* | 0.668 | + | $2.6 \times 10^{-6}$ | + | $3.5 \times 10^{-3}$ |
| 17 | *RP11-707O23.5* | 0.599 | — | $2.8 \times 10^{-6}$ | — | $3.4 \times 10^{-3}$ |
| 17 | *DND1P1* | 0.489 | — | $2.7 \times 10^{-6}$ | — | $3.3 \times 10^{-3}$ |
| 21 | *IFNAR2* | 0.037 | — | $2.2 \times 10^{-11}$ | — | $2.2 \times 10^{-9}$ |

For some of these 11 putative causal genes related to COVID-19 severity, there are already prior knowledge supporting their potential links with COVID-19. For example, SNP *rs1015164* that lies near the antisense transcribed sequence *RP11-24F11*.*2* was associated with HIV set-point viral load [17, 26] and CD4+ T cell counts. Such chemokine receptor-ligand interactions mediating the traffic of inflammatory cells and pathogen-associated immune responses could be plausible for being related to COVID-19 severity. For *FLT1P1*, its expression was reported to be positively associated with predicted neutrophil count [45]. This may mediate the genetic link between this gene and COVID-19 severity. Another identified genes, *CCR5*, is known to play a role in immune cell migration and inflammation. A study found that *CCR5* blockade in critical COVID-19 patients induced decreased inflammatory cytokines, increased CD8 T-cells, and decreased SARS-CoV2 RNA in plasma [29]. For *OAS1*, both predicted and measured protein levels are inversely associated with COVID-19 susceptibility and severity, which is consistent with the current study’s finding [46]. Two of the other genes, namely, *OAS3* and *IFNAR2*, have been identified in our earlier work of COVID-19 TWAS using complementary methods and designs [38].

## 3 Discussion

By leveraging the summary-level expression reference panel with a much larger sample size, our new method SUMMIT substantially improved the prediction accuracy of built expression prediction models, which in turn increased the power for identifying risk genes for complex traits. The corresponding software SUMMIT and its tutorial are available at GitHub.

Through simulations and analyses of GWAS results for 25 traits, we demonstrated the substantial performance gain of SUMMIT over existing methods. Briefly, we demonstrated that SUMMIT improved the expression imputation accuracy (built more expression prediction models with *R*^2^ ≥ 0.01), identified more associations, and achieved higher predictive power for identified ‘silver standard’ genes. Importantly, SUMMIT was applicable to analyze genes with low expression heritability (with *R*^2^ between 0.005 and 0.001), which have larger causal effects sizes on complex traits [41] but have been largely ignored by existing methods.

SUMMIT can be viewed as one type of gene-based Mendelian randomization and provide valid causal interpretations when all genetic variants used in expression prediction models (with non-zero weights) are valid instrumental variables [5, 40, 42]. However, with the wide-spread horizontal pleiotropy of genetic variables [19], valid instrumental variables assumptions may be violated and thus we recommend practitioners using multiple complementary methods jointly to identify likely causal genes. For example, we can apply fine-mapping approaches such FOCUS [23] and FOGS [37] to further prioritize likely causal genes by modeling the linkage disequilibrium and correlation among TWAS signals.

There are several limitations in our current study. First, the summary data of eQTLGen are for whole blood in European ancestry; thus, the built gene expression prediction models would be applicable only for blood tissue of European ancestry subjects. While SUMMIT can be equally applied to other tissues and ancestry, the corresponding summary eQTL data would be needed for such extensions. Second, several TWAS methods such as UTMOST [14] and MR-JTI [44] have been proposed to leverage expression from other tissues or functional annotations to improve the prediction accuracy of expression prediction models. We expect that the number of analyzable genes can be further increased when we integrate this informative information. Third, similar to most existing TWAS methods, the results of SUMMIT imply causality only when valid instrumental variable assumptions are met. A partial solution is to apply fine-mapping to prioritize likely causal genes. However, the robustness of SUMMIT would be significantly improved if we can relax these stringent valid instrumental variable assumptions. We leave this exciting topic to future research.

In conclusion, SUMMIT is a novel and powerful framework to perform TWAS. It integrates summary-level eQTL data with GWAS summary statistics via advanced statistical methods. When combined with fine-mapping and functional validations, its findings may gain insights into the genetic basis of diseases and benefit the development of new therapies. To facilitate such efforts, we deposit TWAS results of 26 traits described in the manuscript into a user-friendly server (https://chongwulab.shinyapps.io/SUMMIT-app/) such that practitioners can search and download gene-trait association results.

## 4 Methods

### 4.1 Penalized regression model for expression prediction

Consider the following linear regression model for estimating the genetically regulated expression:

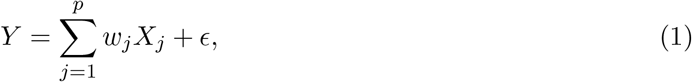

where *Y* is the *N*-dimensional vector of gene expression levels of a gene of interest (corrected for important covariates like age, gender, and genotype principal components),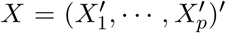 is the *N* × *p* standardized genotype matrix of *p cis*-SNPs around the gene (within 1 MB of the gene transcription start site and transcription end site), the p-dimensional vector *w* = (*w*_1_, …, *w*_*p*_)′ is the *cis*-eQTL effect size, and *ϵ* is random noise with mean zero.

We estimate *w* using a penalized regression framework. Specifically, the objective function is

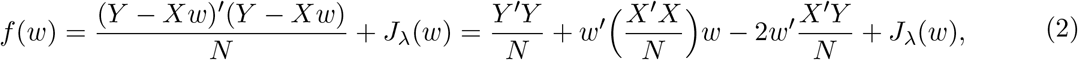

where *J*_*λ*_(·) is a penalty term. Since the performance of different penalties may vary under different genetic architecture, we consider several penalties, including LASSO [32], elastic net [48], minimax concave penalty (MCP) [43], smoothly clipped absolute deviation (SCAD) [7], and MNet [15]. Note that the objective function (Equation (2)) is a function of the marginal statistics *X*′*Y/N* and the linkage disequilirium (LD) matrix *X*′*X/N*, and does not require observing and storing the individual-level data. This allows us to build expression prediction models using eQTL summary-level data, which are computed using a much larger sample size. That is, rewrite the objective function as

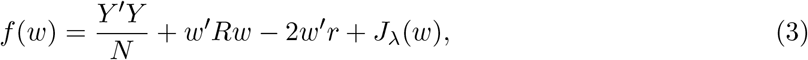

where *r* = *X*′*Y/N* = (*r*_1_, …, *r*_*p*_)′ is a *p*-dimensional vector of standardized marginal effect size for *cis*-SNPs (i.e., correlation between *cis*-SNPs and gene expression levels), and *R* = *X*′*X/N* is the LD matrix of *cis*-SNPs. We use the *z*-scores provided in the summary-level eQTL dataset to estimate *r* (denoted by 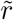) and use a shrinkage estimator (shall be illustrated later) with a LD reference panel (such as 1000 Genomes Project [1]) to estimate *R* (denoted by 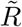). We add an *L*_2_ penalty term *θw*′*w* (where *θ* ≥ 0) to the objective function, which ensures a unique solution upon optimization. Notice that *Y* ′*Y/N* does not depend on *w* and can be ignored when optimizing *f*. Thus, the final objective function that we optimize can be written as,

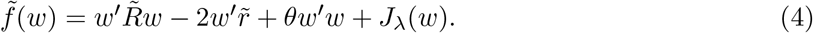

The estimates *ŵ* can be obtained by the coordinate descent algorithm [9], which solves the univariate penalized regression problem sequentially and iteratively. Briefly, suppose that 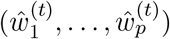 are the coefficients at the *t*-th iteration of the coordinate descent algorithm. Define 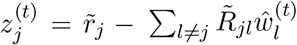.When *J*_*λ*_(*w*) is the LASSO penalty 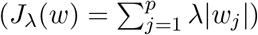, one can update *w*_*j*_ as

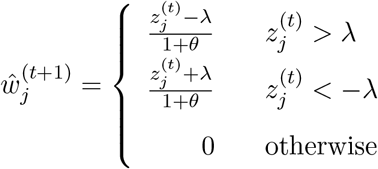

for *j* = 1, …, *p* and *t* = 0, 1, ….

The convergence properties of the coordinate descent algorithm guarantee a local minimum for *ŵ* [9]. We put details on optimization, including choices for initial starting values, *λ*, and *θ* and updating formulas for other penalties to the Supplementary Note.

### 4.2 Estimating standardized marginal effect size 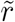 and LD matrix 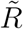

The standardized marginal effect size *r*_*j*_ is often not provided in eQTL summary-level data, but it can be well approximated by 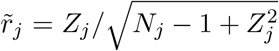, where *Z*_*j*_ and *N*_*j*_ are the *z*-score and sample size for *cis*-SNP *j*, respectively. The eQTL summary-level data combine the results from multiple cohorts and thus the sample size for each SNP may vary. To yield unbiased estimation, we use the SNP-specific sample size *N*_*j*_ instead of the largest sample size (cohort size) [28].

The objective function (4) involves a estimated LD correlation matrix 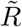. Instead of using the sample correlation matrix estimated from a reference panel such as 1,000 Genomes Project [1] data, we use the shrinkage estimator of the LD matrix [21, 35, 47], which stabilizes the results by shrinking the off-diagonal entries towards zero. Specifically, we first calculate the sample LD correlation matrix from a reference panel. Each entry of the LD correlation matrix is then multiplied by the factor exp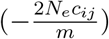, where *N*_*e*_ is the effective population size, *m* is the sample size of the data for generating the genetic map, and *c*_*i*_*j* is the genetic distance between sites *i* and *j* in centiMorgans scale. The entries are set to zero if the factor exp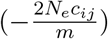 is less than a pre-specified threshold *c*. Following others [21, 47], we use the genetic distance generated from the 1000 Genomes OMNI arrays with *N*_*e*_ = 11, 400 and *m* = 183 and the pre-specified threhold *c* was set to 1 × 10^−3^.

### 4.3 Model training and evaluation

We trained our expression prediction model by using the *cis*-eQTLs summary-level data from eQTLGen [33], which consist of the effect sizes of more than 11 million SNPs from 31, 684 blood samples. Following PrediXcan [10], SNPs in the vicinity of the given gene (within 1 Mbp of the gene transcription start site and transcription end site) were used as the *cis*-genotype information. Further, we filtered out all SNPs with a minor allele frequency < 0.01 and those were non-biallelic or ambiguous or not in HapMap 3 SNPs set [10].

We used both genotype and gene expression data in the GTEx project (version V7, dbGaP Accession number phs000424.v7.p2) [31] to select the tuning parameters. The processed gene expression in whole blood (*N* = 369) were downloaded from the GTEx website. Briefly, the RPKMs in each sample was standardized and normalized by quantile-transformation. Expression for each gene was further adjusted by performing a multivariate linear regression with sex, genotyping platform, 35 PEER factors and three genotype-based principal components (PCs) and the residuals were used as processed expression levels. We used the squared correlation between the predicted and observed expression (that is, *R*^2^) to select the best tuning parameters. Of note, the subjects in GTEx v6 (*N* = 336; 1.1%) have been meta-analyzed in eQTLGen [33] and may result in sub-optimal tuning parameters.

We used independent subjects that are in GTEx v8 but not in GTEx v7 (*N* = 309) as external validation data. Of note, these subjects in GTEx v8 have not been meta-analyzed in eQTLGen and thus can be viewed as an external validation. Because genes with low expression heritability have substantially larger causal effect sizes on complex traits [41], we selected the models with *R*^2^ ≥ 0.005 instead of the commonly used criterion of *R*^2^ ≥ 0.01.

### 4.4 Association Study with single expression prediction model

When individual-level GWAS data (genotype data *X*_new_, phenotype *P*_new_, and covariance matrix *C*_new_) are available, one can apply a generalized linear regression model

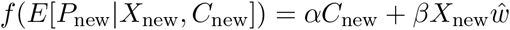

to test *H*_0_ : *β* = 0, where *f* (·) is a link function, and *X*_new_*ŵ* is the predicted genetically regulated expression.

When only summary-level GWAS data are available, one can apply a burden type test:

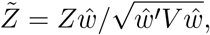

where *Z* is the vector of *z*-scores for all *cis*-SNPs and *V* is the linkage disequilibrium (LD) matrix of analyzed SNPs (which can be estimated by a population reference panel such as 1000 Genomes Project [1]).

### 4.5 Association study with multiple expression prediction models

To further improve the power, we apply the Cauchy combination test [20] to integrate information from *K* models that have *R*^2^ ≥ 0.005. Specifically, we use the following test statistics:

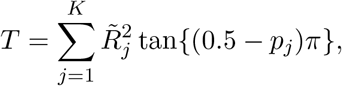

where *p*_*j*_ is the *P*-value for model *j* and 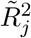 is calculated by 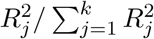. *T* follows a standard Cauchy distribution approximately, and the *P*-value can be calculated as 0.5 − arctan(*T*)*/π*. Of note, we use 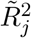 as weights when combining multiple expression prediction models because a larger 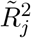 indicates a better expression prediction model. The Cauchy combination test has been widely used in the human genetics community [20, 36], because the *P*-value approximation is accurate for the highly significant results (which are of interest) and there is no need to estimate the correlation structure among the combined *P*-values.

One may be interested in the association direction for a specific gene. For the majority of significant genes identified by SUMMIT, all the expression prediction models yield the same association direction. When expression prediction models provide conflicting association direction, we determine the association direction by the majority voting. In the rare situation where the number of models indicating positive associations equals the number of models indicating negative associations, we declare the association direction is unknown.

### 4.6 Simulation study design

We conducted extensive simulation studies to evaluate how the size of the expression reference panel impacts the expression prediction accuracy and the subsequent power of TWAS. Also, we evaluated whether using the summary-level eQTL data achieves similar performance to that of using individual-level expression reference panel. Specifically, we used data from the UK Biobank (application number 48240) and randomly chose genotype data from 31, 684 (to match the sample size of the eQTLGen data) independent White British individuals as training data, genotype data from additional 369 (to match the sample size of GTEx v7 data) independent White British individuals as tuning data, and genotype data from additional 10, 000 independent White British individuals as test data. The imputed genetic 877 *cis*-SNPs (with minor allele frequency (MAF) > 1%, Hardy-Weinberg p-value > 10^−6^, and imputation “info” score > 0.4) of the arbitrarily chosen gene *CHURC1* were used for our main simulations. We also considered several other randomly selected genes.

We simulated gene expression levels and phenotype values by *E*_*g*_ = *Xw* + *ϵ*_*e*_ and *Y* = *βE*_*g*_ + *ϵ*_*p*_, respectively. *X* is standardized genotype matrix, *w* is the effect size,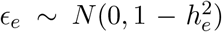, and 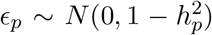, where 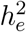 and 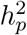 were the expression heritability (i.e., the proportion of gene expression variance explained by SNPs) and phenotypic heritability (i.e., the proportion of phenotypic variance explained by gene expression levels), respectively. We randomly selected *p*_causal_, that is, the proportion of SNPs to be causal, and generated its effect size *w*_*j*_ from *N* (0, 1). The effect sizes for the remaining non-causal SNPs were set to 0. We re-scaled the effect sizes *w* and *β* to achieve the targeted 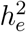 and 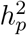.

To evaluate the performance of proposed method SUMMIT, we performed an association scan on the whole simulated training data (*E*_*g*_, *X*) and computed the summary-level data (i.e., *Z* scores) by a linear regression. Besides building models with summary-level data, we also built prediction models with individual-level data of different sample sizes (300, 600, 3, 000, 10, 000, 31, 684). We compared SUMMIT with two widely used methods, PrediXcan [10] and TWAS-fusion [11]. We trained models with PrediXcan and TWAS-fusion with individual-level data of 670 samples (to match the sample size of blood tissue in GTEx v8 data).

We considered comprehensive scenarios that varied the proportion of causal SNPs *p*_causal_ (0.01, 0.05, 0.1, 0.2), expression heritability 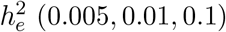, and phenotypic heritability 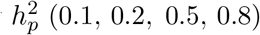. For each scenario, we repeated simulations 1,000 times. Statistical power was calculated as the proportion of 1,000 repeated simulations with *P*-value less than the genome-wide significance threshold 0.05*/*20, 000 = 2.5 × 10^−6^.

### 4.7 Comparison with existing methods

To demonstrate the potential usefulness of the SUMMIT, we further compared SUMMIT with several TWAS methods, including MR-JTI [44], PrediXcan [10], TWAS-fusion [11], and UTMOST [14] for whole blood tissue in the following aspects.

First, we compared prediction accuracy (in terms of *R*^2^) estimated by different methods. Of note, while the prediction performance of competing methods was estimated through cross validations, the prediction performance of SUMMIT was estimated in an external testing dataset. This difference may slightly favor the competing methods. The difference in *R*^2^ across genes was tested by the one-sided Kolmogorov-Smirnov test, a nonparametric test that calculates the largest distance between the empirical distribution functions to determine whether two distributions are equivalent.

Second, we compared different methods by analyzing GWAS summary statistics for 24 complex traits. The details of 24 traits were summarized in Supplementary Table 1. We used the Bonferroni correction for each method with different significance cutoffs as different methods have different numbers of analyzable genes. To make a fair comparison, we also evaluated the common gene set that can be analyzed by all methods and used the same Bonferroni correction cutoff to determine the significant gene sets. The number of significant genes identified by different methods was further compared by Wilcoxon signed-rank test, which compares two matched samples to test whether their population mean ranks differ.

Third, as TWAS can be viewed as a special case of Mendelian randomization [34], we further compared different methods in terms of identifying the causal genes that mediate the association between GWAS loci and the traits of interest. Following [2], we curated a set of likely causal genetrait pairs using information that is independent of GWAS results. Briefly, we utilized the OMIM (Online Mendelian Inheritance in Man) database [13] and rare variant results from exome-wide association studies [18, 22, 24], leading to 1, 287 gene-trait pairs. We used LDetect to partition the genome into approximately independent LD blocks [3] and refined the gene-trait pairs by only considering the genes that were located in LD blocks with at least one genome-wide significant variant, leading to 148 likely causal gene-trait pairs (among 24 distinct traits). We denote the curated set as “silver standard” to highlight their imperfect nature. We compared different methods by the area under receiver operating characteristic curve (AUC).

### 4.8 Applications to COVID-19 GWAS data

To identify novel genes associated with COVID-19 severity, we applied SUMMIT to the GWAS summary data from The COVID-19 HGI (Release 5 (January 2021)) [16]. The detailed information for participating studies, quality control, and analyses have been included on the COVID-19 HGI website (https://www.covid19hg.org/results/). Briefly, data from 9, 986 hospitalized COVID-19 patients and 1, 877, 672 population controls were used in the current analyses. Hospitalized COVID-19 cases represented patients 1) with laboratory confirmed SARS-CoV-2 infection (RNA and/or serology based) and 2) hospitalized due to corona-related symptoms. Controls represent those that are not cases. Only Europeans were included to ensure the homogeneous population structure for the analyses. A fixed-effect meta-analysis of individual participating studies was performed and variants with imputation quality > 0.6 were retained.

We applied fine-mapping method FOGS [37] to prioritize likely causal genes for COVID-19 severity. In the end, we evaluated associations of identified genes with additional COVID-19 phenotype. Briefly, we leveraged A2 ALL eur (Europeans; 5, 101 cases and 1, 383, 241 controls) for comparing very severe respiratory confirmed COVID vs population controls.

## Data Availability

All data produced in the present work are either contained in the manuscript or available online at https://chongwulab.shinyapps.io/SUMMIT-app/.

## Data availability statement

The GWAS summary data (with download link) used in this study are summarized in Supplementary Table 1. The eQTL summary data are available at https://www.eqtlgen.org/cis-eqtls.html. The UK Biobank is an open-access resource, available at https://www.ukbiobank.ac.uk/researchers/. This research was conducted with approved access to UK Biobank data under application numbers 48240. The genotype and RNA sequencing data for the GTEx project are available at the database of Genotypes and Phenotypes (accession number phs000424.v8.p2). The processed gene expression for the GTEx project is available from the GTEx portal (https://gtexportal.org). The SUMMIT models will be available for free download from Zenodo. All real data results are available at https://chongwulab.shinyapps.io/SUMMIT-app/, where practitioners can search and download results easily.

## Codes availability statement

The software SUMMIT and the code for reproducing the results described in this study will be available from GitHub (https://github.com/ChongWuLab/SUMMIT).

## Acknowledgement

This study is supported by National Institutes of Health (R03 AG070669) and has been conducted using the UK Biobank recourse (application numbers 48240). The authors would like to thank all of the individuals for their participation in GWAS studies and UK Biobank and all the researchers, clinicians, technicians and administrative staff for their contribution to the studies and making their GWAS summary results publicly available.

## References

[1] 1000 Genomes Project Consortium (2015). A global reference for human genetic variation. Nature, 526(7571):68–74.

[2] Barbeira, A. N., Bonazzola, R., Gamazon, E. R., Liang, Y., Park, Y., Kim-Hellmuth, S., Wang, G., Jiang, Z., Zhou, D., Hormozdiari, F., et al. (2021). Exploiting the gtex resources to decipher the mechanisms at gwas loci. Genome Biology, 22(1):1–24.

[3] Berisa, T. and Pickrell, J. K. (2016). Approximately independent linkage disequilibrium blocks in human populations. Bioinformatics, 32(2):283–285.

[4] Boyle, E. A., Li, Y. I., and Pritchard, J. K. (2017). An expanded view of complex traits: from polygenic to omnigenic. Cell, 169(7):1177–1186.

[5] Burgess, S. and Thompson, S. G. (2013). Use of allele scores as instrumental variables for mendelian randomization. International Journal of Epidemiology, 42(4):1134–1144.

[6] Consortium, G. et al. (2020). The gtex consortium atlas of genetic regulatory effects across human tissues. Science, 369(6509):1318–1330.

[7] Fan, J. and Li, R. (2001). Variable selection via nonconcave penalized likelihood and its oracle properties. Journal of the American statistical Association, 96(456):1348–1360.

[8] Finucane, H. K., Bulik-Sullivan, B., Gusev, A., Trynka, G., Reshef, Y., Loh, P.-R., Anttila, V., Xu, H., Zang, C., Farh, K., et al. (2015). Partitioning heritability by functional annotation using genome-wide association summary statistics. Nature Genetics, 47(11):1228.

[9] Friedman, J., Hastie, T., and Tibshirani, R. (2010). Regularization paths for generalized linear models via coordinate descent. Journal of Statistical Software, 33(1):1.

[10] Gamazon, E. R., Wheeler, H. E., Shah, K. P., Mozaffari, S. V., Aquino-Michaels, K., Carroll, R. J., Eyler, A. E., Denny, J. C., Nicolae, D. L., Cox, N. J., et al. (2015). A gene-based association method for mapping traits using reference transcriptome data. Nature Genetics, 47(9):1091–1098.

[11] Gusev, A., Ko, A., Shi, H., Bhatia, G., Chung, W., Penninx, B. W., Jansen, R., De Geus, E. J., Boomsma, D. I., Wright, F. A., et al. (2016). Integrative approaches for large-scale transcriptome-wide association studies. Nature Genetics, 48(3):245–252.

[12] Gusev, A., Mancuso, N., Won, H., Kousi, M., Finucane, H. K., Reshef, Y., Song, L., Safi, A., McCarroll, S., Neale, B. M., et al. (2018). Transcriptome-wide association study of schizophrenia and chromatin activity yields mechanistic disease insights. Nature Genetics, 50(4):538–548.

[13] Hamosh, A., Scott, A. F., Amberger, J. S., Bocchini, C. A., and McKusick, V. A. (2005). Online mendelian inheritance in man (omim), a knowledgebase of human genes and genetic disorders. Nucleic Acids Research, 33(suppl 1):D514–D517.

[14] Hu, Y., Li, M., Lu, Q., Weng, H., Wang, J., Zekavat, S. M., Yu, Z., Li, B., Gu, J., Muchnik, S., et al. (2019). A statistical framework for cross-tissue transcriptome-wide association analysis. Nature Genetics, 51(3):568–576.

[15] Huang, J., Breheny, P., Lee, S., Ma, S., and Zhang, C.-H. (2016). The mnet method for variable selection. Statistica Sinica, pages 903–923.

[16] Initiative, C.-. H. G. et al. (2021). Mapping the human genetic architecture of covid-19 by worldwide meta-analysis. MedRxiv.

[17] Kulkarni, S., Lied, A., Kulkarni, V., Rucevic, M., Martin, M. P., Walker-Sperling, V., Anderson, S. K., Ewy, R., Singh, S., Nguyen, H., et al. (2019). Ccr5as lncrna variation differentially regulates ccr5, influencing hiv disease outcome. Nature Immunology, 20(7):824–834.

[18] Liu, D. J., Peloso, G. M., Yu, H., Butterworth, A. S., Wang, X., Mahajan, A., Saleheen, D., Emdin, C., Alam, D., Alves, A. C., et al. (2017). Exome-wide association study of plasma lipids in¿ 300,000 individuals. Nature Genetics, 49(12):1758–1766.

[19] Liu, X., Li, Y. I., and Pritchard, J. K. (2019a). Trans effects on gene expression can drive omnigenic inheritance. Cell, 177(4):1022–1034.

[20] Liu, Y., Chen, S., Li, Z., Morrison, A. C., Boerwinkle, E., and Lin, X. (2019b). ACAT: A fast and powerful p value combination method for rare-variant analysis in sequencing studies. The American Journal of Human Genetics, 104(3):410–421.

[21] Lloyd-Jones, L. R., Zeng, J., Sidorenko, J., Yengo, L., Moser, G., Kemper, K. E., Wang, H., Zheng, Z., Magi, R., Esko, T., et al. (2019). Improved polygenic prediction by bayesian multiple regression on summary statistics. Nature Communications, 10(1):1–11.

[22] Locke, A. E., Steinberg, K. M., Chiang, C. W., Service, S. K., Havulinna, A. S., Stell, L., Pirinen, M., Abel, H. J., Chiang, C. C., Fulton, R. S., et al. (2019). Exome sequencing of finnish isolates enhances rare-variant association power. Nature, 572(7769):323–328.

[23] Mancuso, N., Freund, M. K., Johnson, R., Shi, H., Kichaev, G., Gusev, A., and Pasaniuc, B. (2019). Probabilistic fine-mapping of transcriptome-wide association studies. Nature Genetics, 51(4):675–682.

[24] Marouli, E., Graff, M., Medina-Gomez, C., Lo, K. S., Wood, A. R., Kjaer, T. R., Fine, R. S., Lu, Y., Schurmann, C., Highland, H. M., et al. (2017). Rare and low-frequency coding variants alter human adult height. Nature, 542(7640):186–190.

[25] Maurano, M. T., Humbert, R., Rynes, E., Thurman, R. E., Haugen, E., Wang, H., Reynolds, A. P., Sandstrom, R., Qu, H., Brody, J., et al. (2012). Systematic localization of common disease-associated variation in regulatory dna. Science, 337(6099):1190–1195.

[26] McLaren, P. J., Coulonges, C., Bartha, I., Lenz, T. L., Deutsch, A. J., Bashirova, A., Buchbinder, S., Carrington, M. N., Cossarizza, A., Dalmau, J., et al. (2015). Polymorphisms of large effect explain the majority of the host genetic contribution to variation of hiv-1 virus load. Proceedings of the National Academy of Sciences, 112(47):14658–14663.

[27] Nagpal, S., Meng, X., Epstein, M. P., Tsoi, L. C., Patrick, M., Gibson, G., De Jager, P. L., Bennett, D. A., Wingo, A. P., Wingo, T. S., et al. (2019). Tigar: An improved bayesian tool for transcriptomic data imputation enhances gene mapping of complex traits. The American Journal of Human Genetics, 105(2):258–266.

[28] Palmer, C. and Peer, I. (2017). Statistical correction of the winner’s curse explains replication variability in quantitative trait genome-wide association studies. PLOS Genetics, 13(7):e1006916.

[29] Patterson, B. K., Seethamraju, H., Dhody, K., Corley, M. J., Kazempour, K., Lalezari, J., Pang, A. P., Sugai, C., Mahyari, E., Francisco, E. B., et al. (2021). Ccr5 inhibition in critical covid-19 patients decreases inflammatory cytokines, increases cd8 t-cells, and decreases sars-cov2 rna in plasma by day 14. International Journal of Infectious Diseases, 103:25–32.

[30] Raj, T., Li, Y. I., Wong, G., Humphrey, J., Wang, M., Ramdhani, S., Wang, Y.-C., Ng, B., Gupta, I., Haroutunian, V., et al. (2018). Integrative transcriptome analyses of the aging brain implicate altered splicing in alzheimer’s disease susceptibility. Nature Genetics, 50(11):1584–1592.

[31] GTEx Consortium (2017). Genetic effects on gene expression across human tissues. Nature, 550(7675):204–213.

[32] Tibshirani, R. (1996). Regression shrinkage and selection via the lasso. Journal of the Royal Statistical Society: Series B (Methodological), 58(1):267–288.

[33] V∼osa, U., Claringbould, A., Westra, H.-J., Bonder, M. J., Deelen, P., Zeng, B., Kirsten, H., Saha, A., Kreuzhuber, R., Kasela, S., et al. (2018). Unraveling the polygenic architecture of complex traits using blood eqtl meta-analysis. bioRxiv, page 447367.

[34] Wainberg, M., Sinnott-Armstrong, N., Mancuso, N., Barbeira, A. N., Knowles, D. A., Golan, D., Ermel, R., Ruusalepp, A., Quertermous, T., Hao, K., et al. (2019). Opportunities and challenges for transcriptome-wide association studies. Nature Genetics, 51(4):592–599.

[35] Wen, X. and Stephens, M. (2010). Using linear predictors to impute allele frequencies from summary or pooled genotype data. The Annals of Applied Statistics, 4(3):1158.

[36] Wu, C., Bradley, J., Li, Y., Wu, L., and Deng, H.-w. (2021a). A gene-level methylome-wide association analysis identifies novel alzheimer’s disease genes. Bioinformatics.

[37] Wu, C. and Pan, W. (2020). A powerful fine-mapping method for transcriptome-wide association studies. Human genetics, 139(2):199–213.

[38] Wu, L., Zhu, J., Liu, D., Sun, Y., and Wu, C. (2021b). An integrative multiomics analysis identifies putative causal genes for covid-19 severity. Genetics in Medicine, pages 1–11.

[39] Xu, Z., Wu, C., Wei, P., and Pan, W. (2017). A powerful framework for integrating eQTL and GWAS summary data. Genetics, 207:893–902.

[40] Xue, H., Pan, W., and Initiative, A. D. N. (2020). Some statistical consideration in transcriptome-wide association studies. Genetic Epidemiology, 44(3):221–232.

[41] Yao, D. W., O’Connor, L. J., Price, A. L., and Gusev, A. (2020). Quantifying genetic effects on disease mediated by assayed gene expression levels. Nature Genetics, pages 1–8.

[42] Yuan, Z., Zhu, H., Zeng, P., Yang, S., Sun, S., Yang, C., Liu, J., and Zhou, X. (2020). Testing and controlling for horizontal pleiotropy with probabilistic mendelian randomization in transcriptome-wide association studies. Nature Communications, 11(1):1–14.

[43] Zhang, C.-H. et al. (2010). Nearly unbiased variable selection under minimax concave penalty. The Annals of Statistics, 38(2):894–942.

[44] Zhou, D., Jiang, Y., Zhong, X., Cox, N. J., Liu, C., and Gamazon, E. R. (2020). A unified framework for joint-tissue transcriptome-wide association and mendelian randomization analysis. Nature Genetics, 52(11):1239–1246.

[45] Zhou, J., Sun, Y., Huang, W., and Ye, K. (2021a). Altered blood cell traits underlie a major genetic locus of severe covid-19. The Journals of Gerontology: Series A, 76(8):e147–e154.

[46] Zhou, S., Butler-Laporte, G., Nakanishi, T., Morrison, D. R., Afilalo, J., Afilalo, M., Laurent, L., Pietzner, M., Kerrison, N., Zhao, K., et al. (2021b). A neanderthal oas1 isoform protects individuals of european ancestry against covid-19 susceptibility and severity. Nature Medicine, 27(4):659–667.

[47] Zhu, X. and Stephens, M. (2017). Bayesian large-scale multiple regression with summary statistics from genome-wide association studies. The Annals of Applied Statistics, 11(3):1561.

[48] Zou, H. and Hastie, T. (2005). Regularization and variable selection via the elastic net. Journal of the Royal Statistical Society: Series B, 67(2):301–320.

