## Supplementary for "SUMMIT: An integrative approach for better transcriptomic data imputation improves causal gene identification"

#### 1 Details on the optimization of SUMMIT

We start with the objective function $\tilde{f}(w)$,

$$\tilde{f}(w) = w'\tilde{R}w - 2w'\tilde{r} + \theta w'w + J_\lambda(w),$$

where $w$ is the effect sizes of $p$ SNPs, $\tilde{R}$ is the estimated covariance matrix, $\tilde{r}$ is the estimated correlation between *cis*-SNPs and the gene expression level, $\theta$ is the tuning parameter of $L_2$ regularization, $J_\lambda(w)$ is a to-be-determined penalty term with tuning parameter $\lambda$.

The objective function above be solved by the coordinate descent algorithm, which sequentially and iteratively solves the univariate penalized regression problem. Assume that $(\hat{w}_1^{(t)}, \ldots, \hat{w}_p^{(t)})$ are

the coefficients at iteration $t$ and we define

$$z_j^{(t)} = \tilde{r}_j - \sum_{l \neq j} \tilde{R}_{jl} \hat{w}_l^{(t)}.$$

By solving the univariate penalized regression problem (with $J_\lambda(w)$) given current estimates $(\hat{w}_1^{(t)}, \ldots, \hat{w}_p^{(t)})$, one can update $w_j$ accordingly. In the following, we will briefly describe the types of penalty we used and their corresponding updating formula.

**LASSO and Elastic Net**

When using the LASSO (least absolute shrinkage and selection operator) [3] and Elastic Net [5], the penalty term for $w_j$ can be written as,

$$J_\lambda(w_j) = 2\lambda((1-\alpha)|w_j| + \alpha w_j^2),$$

where $\alpha$ is a hyperparameter that controls the ratio of $L_1$ and $L_2$ penalty. LASSO can be viewed as a special case of Elastic Net with $\alpha = 0$. For Elastic Net, we set $\alpha = 0.5$.

The updating formula for $w_j$ is,

$$\hat{w}_j^{(t+1)} = \frac{\mathcal{S}(z_j^{(t)}, \lambda(1-\alpha))}{1 + \theta + 2\alpha\lambda},$$

where $\mathcal{S}(U, \lambda)$ is the soft-thresholding operator, which is defined as,

$$\mathcal{S}(U, \lambda) = \begin{cases} U - \lambda, & U > \lambda; \\ U + \lambda, & U < -\lambda; \\ 0, & \text{otherwise.} \end{cases}$$

**MCP and MNet**

When using the MCP (Minimax concave penalty) [4], the penalty term for $w_j$ can be written as,

$$J_\lambda(w_j) = \begin{cases} 2(\lambda|w_j| - \frac{w_j^2}{2a}), & |w_j| \leq a\lambda; \\ a\lambda^2, & \text{otherwise.} \end{cases}$$

The updating formula for $w_j$ is,

$$\hat{w}_j^{(t+1)} = \begin{cases} \frac{\mathcal{S}(z_j^{(t)},\lambda)}{1+\theta-\frac{1}{a}}, & |w_j| \leq a\lambda; \\ \frac{z_j^{(t)}}{1+\theta}, & \text{otherwise.} \end{cases}$$

When using the MNet [2], the penalty term for $w_j$ can be written as,

$$J_\lambda(w_j) = \begin{cases} 2\alpha(\lambda|w_j| - \frac{w_j^2}{2a}) + 2(1-\alpha)w_j^2, & |w_j| \leq a(\alpha\lambda)(1+(1-\alpha)\lambda); \\ a\alpha\lambda^2 + 2(1-\alpha)w_j^2, & \text{otherwise.} \end{cases}$$

The updating formula for $w_j$ is,

$$\hat{w}_j^{(t+1)} = \begin{cases} \frac{\mathcal{S}(z_j^{(t)},\alpha\lambda)}{1+\theta+(1-\alpha)-\frac{1}{a}}, & |w_j| \leq a(\alpha\lambda)(1+(1-\alpha)\lambda); \\ \frac{z_j^{(t)}}{1+\theta+(1-\alpha)\lambda}, & \text{otherwise.} \end{cases}$$

For both MCP and MNet, we set $a = 3$. For MNet, we set $\alpha = 0.5$.

**SCAD**

When using the SCAD (Smoothly clipped absolute deviation) [1], the penalty term for $w_j$ can be written as,

$$J_\lambda(w_j) = \begin{cases} 2(\lambda|w_j|), & |w_j| \leq \lambda; \\ \frac{2a\lambda|w_j|-w_j^2-\lambda^2}{a-1}, & \lambda < |w_j| \leq a\lambda; \\ \lambda^2(a+1), & \text{otherwise.} \end{cases}$$

The corresponding updating formula for $w_j$ is

$$\hat{w}_j^{(t+1)} = \begin{cases} \frac{\mathcal{S}(z_j^{(t)},\lambda)}{1+\theta}, & |w_j| \leq \lambda; \\ \frac{\mathcal{S}(z_j^{(t)},\frac{a}{a-1}\lambda)}{1+\theta-\frac{1}{a-1}}, & \lambda < |w_j| \leq a\lambda; \\ \frac{z_j^{(t)}}{1+\theta}, & \text{otherwise}. \end{cases}$$

For SCAD, we set $a = 3.7$.

##### Search space of tuning parameters

We apply warm start to generate a solution path for $\lambda$ and search $\theta$ in the set of $(0.1, 0.2, \ldots, 0.9)$. The "optimal" tuning parameters are selected based on maximizing $R^2$ in a tuning dataset.

### 2 Additional simulation figures

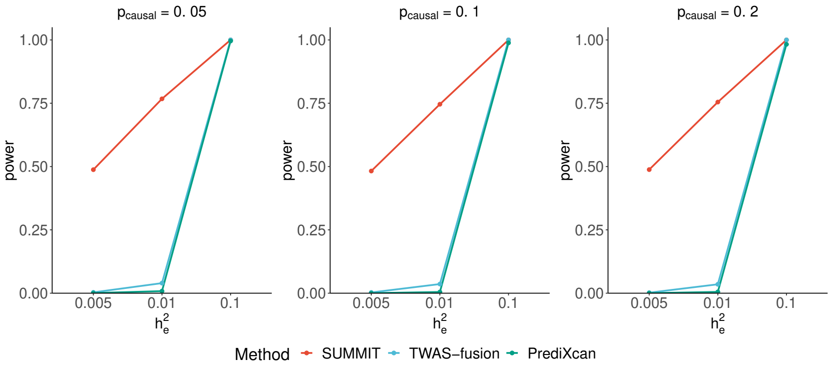


(a) $h_p^2 = 0.1$

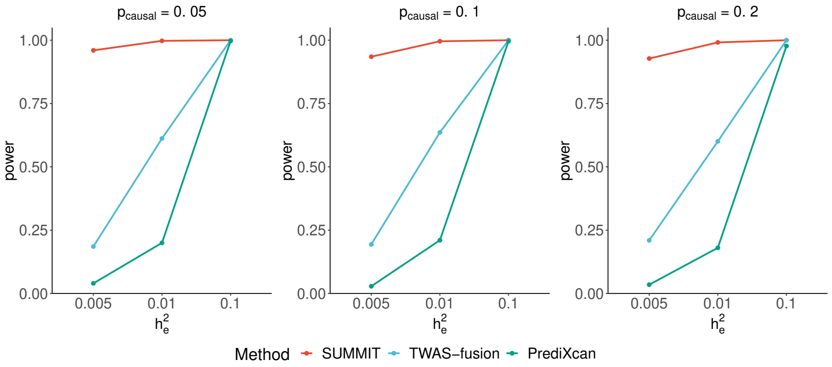


(b) $h_p^2 = 0.5$

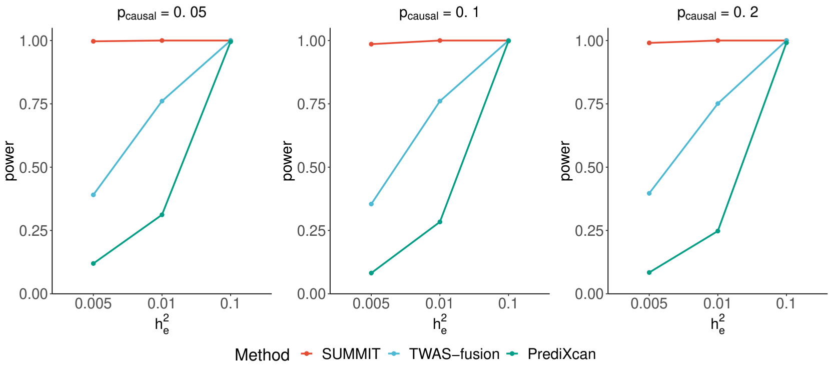


(c) $h_p^2 = 0.8$

Figure S1: **Empirical power comparisons with different $h_p^2$ based on gene *CHURC1*.** The empirical power was estimated by the proportions of $P$-value less than the significance threshold $2.5 \times 10^{-6}$.

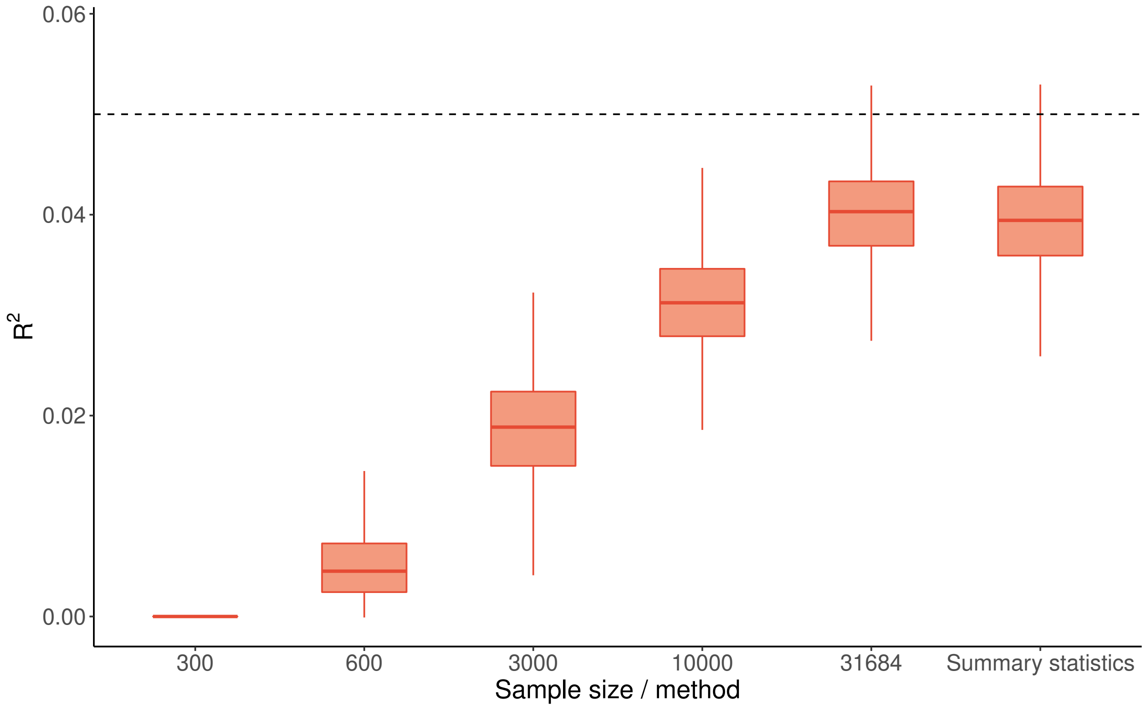


Figure S2: **Increasing training sample size increases $R^2$ and SUMMIT achieves the results similar to those of using individual-level data for simulations based on gene *CHURC1*.** We set $h_e^2 = 0.05$ and $p_{causal} = 0.2$. $R^2$ was calculated in a testing dataset. The "Summary statistics" represents applying SUMMIT with summary-level data.

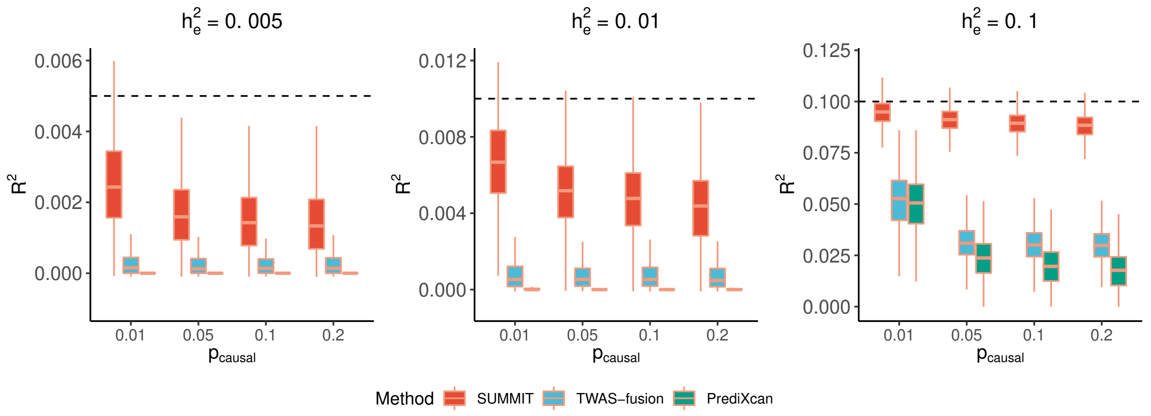


(a) Imputation $R^2$

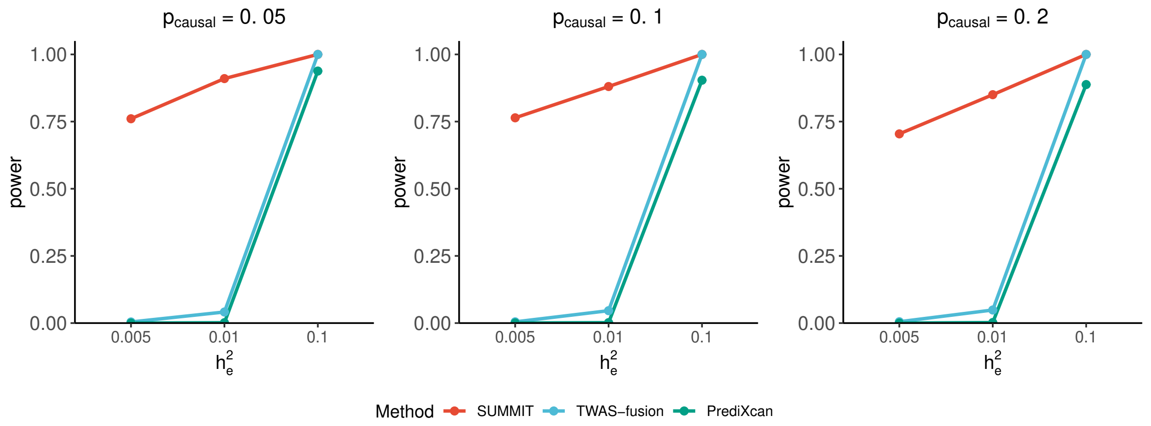


(b) Statistical power

Figure S3: **Performance comparison in simulations based on gene *ICA1*.** Plots of imputation $R^2$ (a) and subsequent power (b) in test samples by SUMMIT, TWAS-fusion, and PrediXcan, with varying true expression heritability $h_e^2$ and proportion of true causal SNPs $p_{\text{causal}}$. For (b), we set $h_p^2 = 0.2$ and empirical power was estimated by the proportions of $P$-value less than the significance threshold $2.5 \times 10^{-6}$.

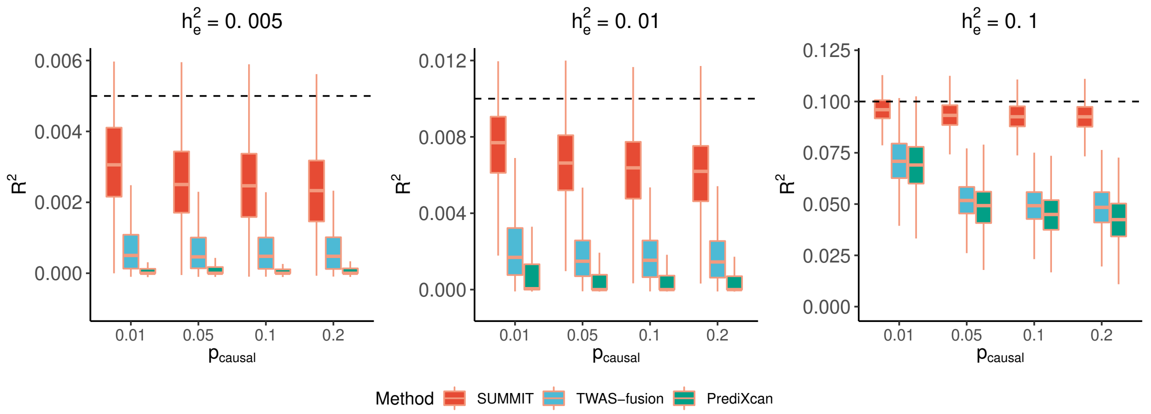


(a) Imputation $R^2$

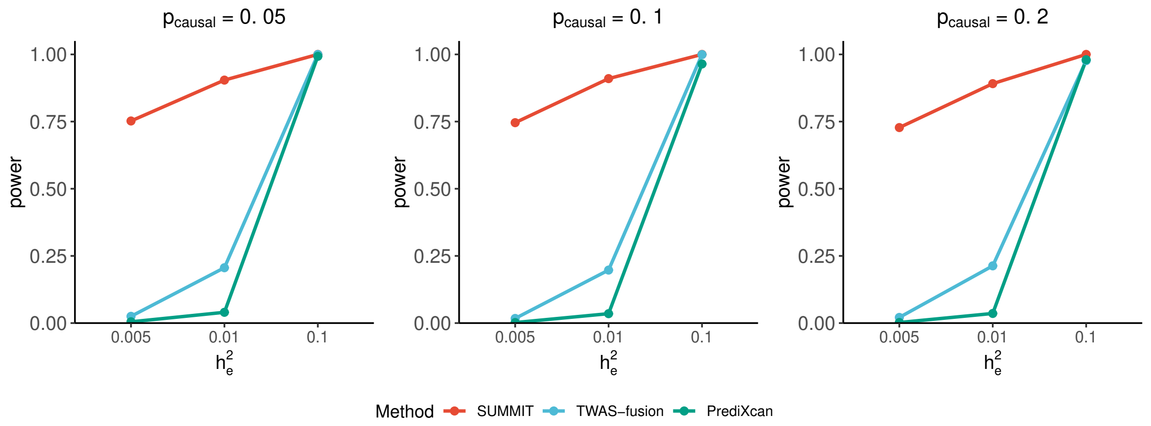


(b) Statistical power

Figure S4: **Performance comparison in simulations based on gene *KRIT1*.** Plots of imputation $R^2$ (a) and subsequent power (b) in test samples by SUMMIT, TWAS-fusion, and PrediXcan, with varying true expression heritability $h_e^2$ and proportion of true causal SNPs $p_{\text{causal}}$. For (b), we set $h_p^2 = 0.2$ and empirical power was estimated by the proportions of $P$-value less than the significance threshold $2.5 \times 10^{-6}$.

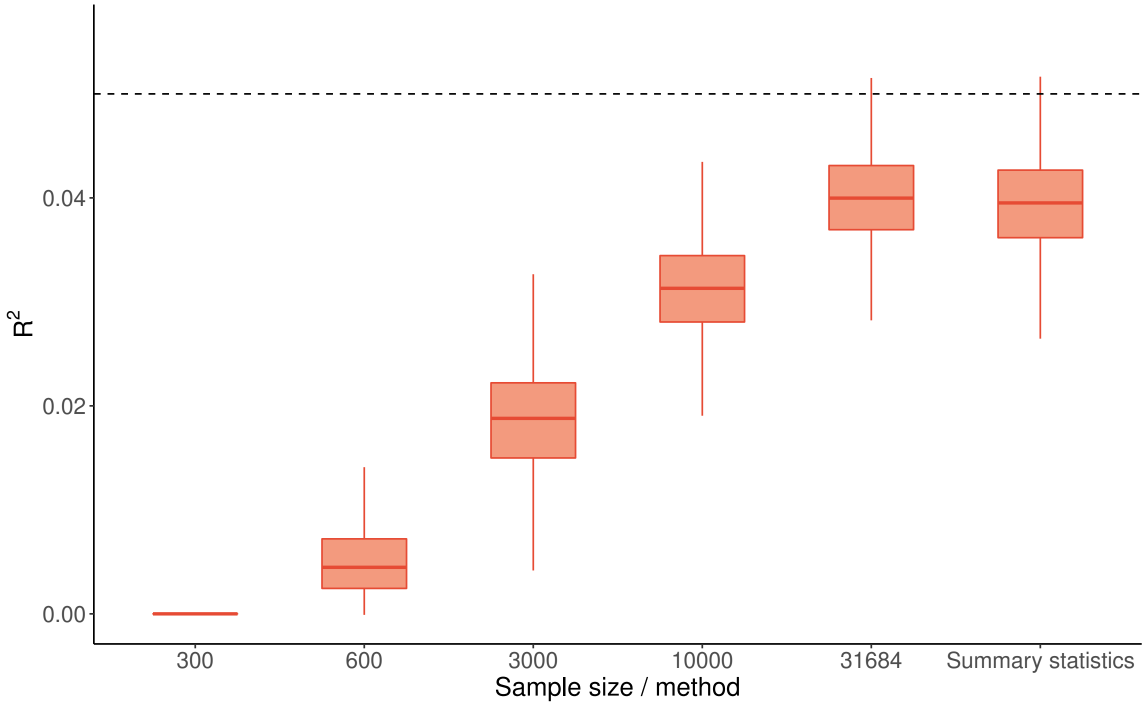


Figure S5: **Distribution of imputation $R^2$ for gene *ICA1* with respect to different sample sizes, with $h_e^2 = 0.05$ and $p_{causal} = 0.2$.** The "Summary statistics" bar was based on simulations using summary-level data to estimate $\tilde{r}$.

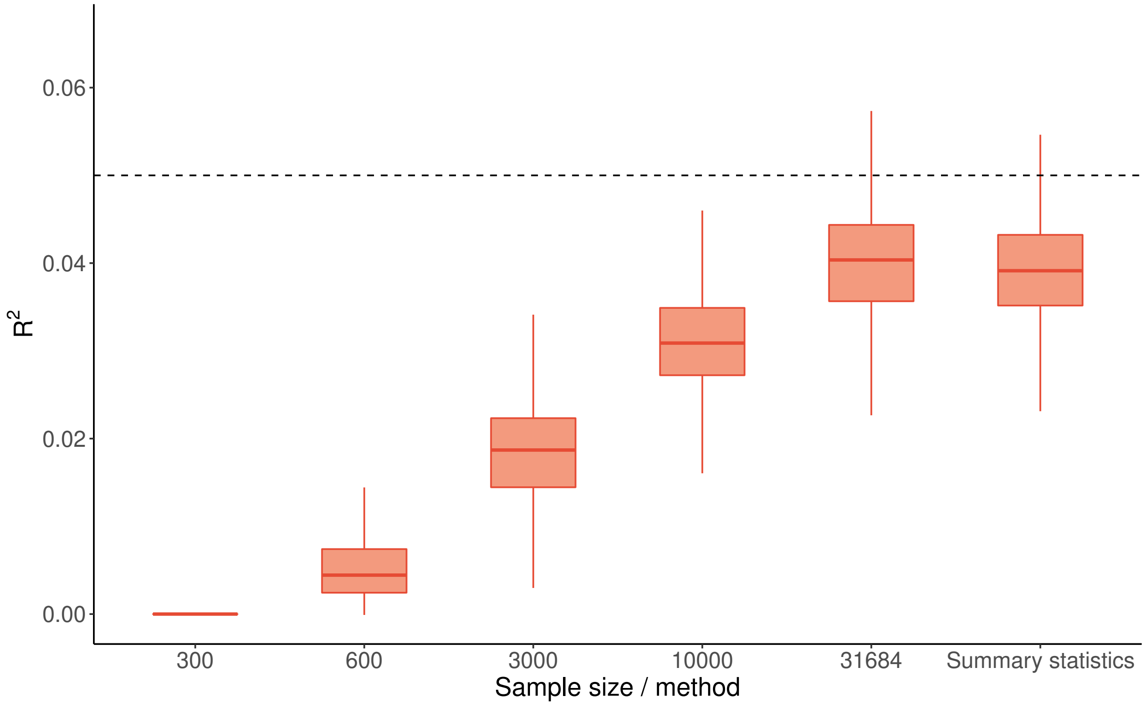


Figure S6: **Distribution of imputation $R^2$ for gene *KRIT1* with respect to different sample sizes, with $h_e^2 = 0.05$ and $p_{causal} = 0.2$.** The "Summary statistics" bar was based on simulations using summary-level data to estimate $\tilde{r}$.

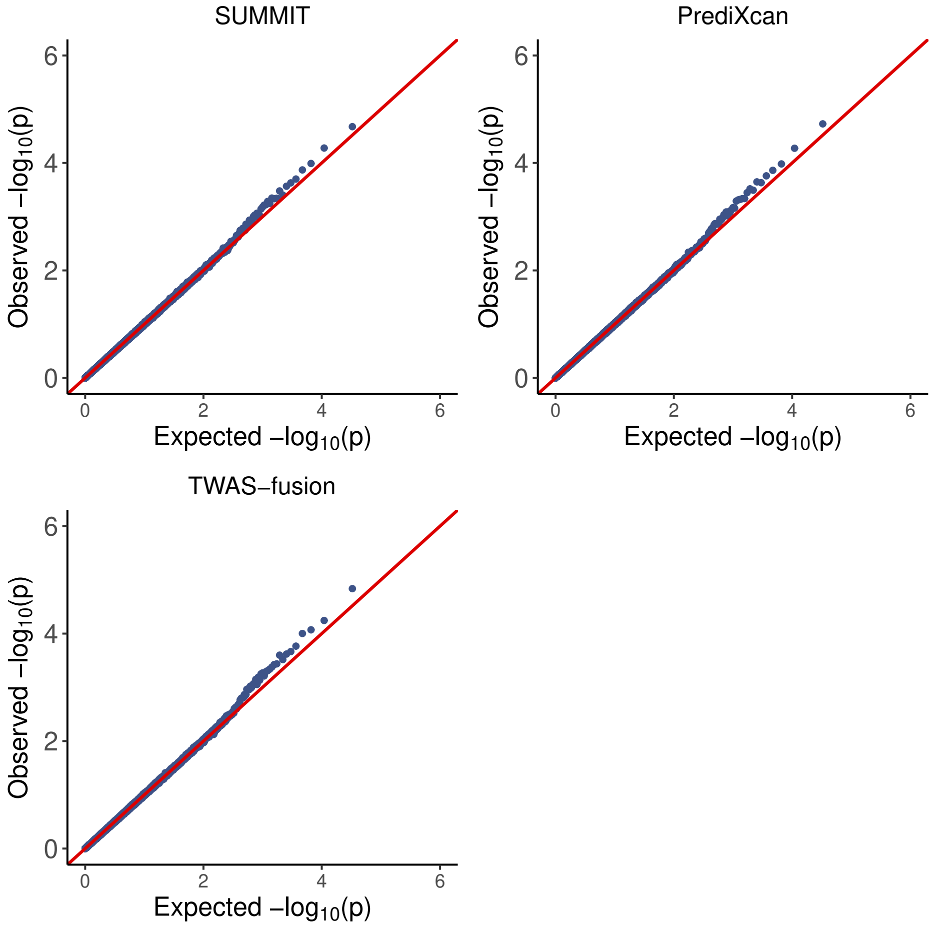


Figure S7: **QQ plots of $P$-values under the null hypothesis based on gene *CHURC1*.** We ran 5,000,000 simulations (5,000 runs for each of 1,000 computed weights) under the null hypothesis to evaluate the Type 1 error rates.

### References


[1] Fan, J. and Li, R. (2001). Variable selection via nonconcave penalized likelihood and its oracle properties. *Journal of the American statistical Association*, 96(456):1348–1360.

[2] Huang, J., Breheny, P., Lee, S., Ma, S., and Zhang, C.-H. (2016). The mnet method for variable selection. *Statistica Sinica*, pages 903–923.

[3] Tibshirani, R. (1996). Regression shrinkage and selection via the lasso. *Journal of the Royal Statistical Society: Series B (Methodological)*, 58(1):267–288.

[4] Zhang, C.-H. et al. (2010). Nearly unbiased variable selection under minimax concave penalty. *The Annals of Statistics*, 38(2):894–942.

[5] Zou, H. and Hastie, T. (2005). Regularization and variable selection via the elastic net. *Journal of the Royal Statistical Society: Series B*, 67(2):301–320.
